# First phylogenetic analysis of Malian SARS-CoV-2 sequences provide molecular insights into the genomic diversity of the Sahel region

**DOI:** 10.1101/2020.09.23.20165639

**Authors:** B Kouriba, A Dürr, A Rehn, AK Sangaré, BY Traoré, MS Bestehorn-Willmann, WLJ Ouedraogo, A Heitzer, E Sogodogo, A Maiga, MC Walter, F Zimmermann, R Wölfel, MH Antwerpen

## Abstract

We are currently facing a pandemic of COVID-19, caused by a spillover from an animal-originating coronavirus to humans occuring in the Wuhan region, China, in December 2019. From China the virus has spread to 188 countries and regions worldwide, reaching the Sahel region on the 2nd of March 2020. Since whole genome sequencing (WGS) data is very crucial to understand the spreading dynamics of the ongoing pandemic, but only limited sequence data is available from the Sahel region to date, we have focused our efforts on generating the first Malian sequencing data available. Screening of 217 Malian patient samples for the presence of SARS-CoV-2 resulted in 38 positive isolates from which 21 whole genome sequences were generated. Our analysis shows that both, the early A (19B) and the fast evolving B (20A/C) clade, are present in Mali indicating multiple and independent introductions of the SARS-CoV-2 to the Sahel region.

## Introduction

SARS-CoV-2 (previously called 2019-nCoV) is a novel member in the genus of Betacoronaviridae and is responsible for the current, rapidly escalating COVID-19 pandemic. To this day, this virus, which has a lower pathogenicity than SARS-CoV but a higher human to human transmissibility [1], caused more than 16 million infections and 648,000 associated deaths worldwide ([2] 27 July 2020)

The outbreak of the virus began in China, in the city of Wuhan, in December 2019, followed by the shift of the epicenter to Europe in mid-March 2020 [3]. From Europe and Asia, the wave of infection moved to North and South America and as well to Africa. The first COVID-19 case in the Sahel region was reported from Senegal on the 2nd of March 2020 [4]. Within 3 weeks, the virus did spread to all Sahel countries, including Burkina Faso, Mauritania, Tchad, Niger, Sudan and Eritrea, reaching Mali on the 25th of March 2020 [5]; [6]. Since the Sahel region is currently facing several challenges, including a severe food and security crisis, it is difficult to judge the effects, the COVID-19 pandemic will have on the already overwhelmed healthcare systems [7]. Especially in conflict-affected areas where the population has only limited access to clean drinking water and handwashing facilities, hundreds of health centres are closed or are not operating due to the poor security situation. Furthermore, social distancing measures are difficult to implement in the dense African urban settings and the national governments struggle to divide the already insufficient budget between health, food and security emergencies [8]. Therefore it is not surprising that the medical and technical equipment as well as the laboratory infrastructure in those countries is sometimes outdated and not on a comparable level to most European countries or the USA. Hence, only limited genomic data of SARS-CoV-2 exists from the Sahel region, with no whole genome sequences being available from Mali to this date.

On 25.03.2020, the first two COVID-19 cases (a 49-year-old woman living in Bamako and a 62-year-old patient from Kayes, both returning from France) were confirmed in Mali [6]. To support the Malian public health system, SARS-CoV-2 diagnostic capabilities were established at the *Centre d’Infectiologie Charles Mérieux du Mali* (CICM-Mali) by training of scientific personnel and by providing testing reagents supported by the German Enhancement Initiative against Biological Threats in the G5 Sahel region. Based on the given training, the CICM-Mali was prepared as the second and central diagnostic center to support the diagnosis of SARS-CoV-2 for Bamako and its surrounding regions and started its activities on April 3rd.

Since the confirmation of the first two cases in March 2020, a steady increase in the number of total cases and a fast spreading of the SARS-CoV-2 virus within Mali was observed. As of 26 July 2020, 2510 cumulative COVID-19 cases and 123 (Case fatality ratio: 4,93%) related deaths have been reported in Mali. [11] Based on the current data available, 13 out of 100 000 Malians get infected resulting in a moderate risk for the Malian population to acquire COVID-19, especially considering the fact that mitigation strategies, like social distancing, cannot be applied to the same extent as in Europe [9]. In order to analyse origins of existing infections, distribution patterns and hence limit the spread of SARS-CoV-2, sequencing data of COVID-19 positive cases are needed.

In this study, we analyzed the first Malian genome sequences of SARS CoV-2 originating from patients from Bamako and its surrounding villages. Using comparative genomics we set the results in the context of available African genome sequences thereby providing data of underrepresented regions, like the Sahel, to the scientific community.

## Material and Methods

### Patient sampling

Nasopharyngeal or oropharyngeal samples were collected from 217 suspected patients using FLOQ swabs (Copan Diagnostics, Murrieta, USA). Afterwards, the swabs were stored in UTM transport medium (Copan Diagnostics, Murrieta, USA). Sampling was performed at the reference health centers of the affected health districts of Bamako, the capital city of Mali, of the regions of Kayes, Koulikoro, Segou and Mopti. Samples were first collected at the National Institute of Public Health then transferred to the *Centre d’Infectiologie Charles Mérieux du Mali* for analysis.

### RNA extraction and SARS-CoV-2 detection

Viral RNA was extracted using the QIAamp Viral RNA Mini Kit (Qiagen, Hilden, Germany) and analyzed for the presence of SARS-CoV-2 at the *Centre d’Infectiologie Charles Mérieux du Mali* by reverse transcription quantitative PCR (RT-qPCR) according to the published protocol of [10] targeting the RdRp and E gene region. SuperScript III Platinum Polymerase qRT-PCR kit (ThermoFisher Scientific, Germering, Germany) was used for amplification. MS2 phages were added as internal control. In order to allow simultaneous detection of the E and MS2 gene, the E singleplex assay of Corman et al. was converted into a multiplex assay by addition of MS2 specific primers and probes (Cy5-labeled) [12]. Positive results obtained in the E gene assay were confirmed using the discriminatory RT-qPCR RdRp gene assay.

38 samples which had been tested positive for both, the E and the RdRp gene, were sent to the Bundeswehr Institute of Microbiology (IMB) for whole genome sequencing.

### Library preparation and sequencing

All samples were processed according to the published nCoV-2019 ARTIC sequencing protocol [13,14]. To this end, cDNA Synthesis of extracted RNA was performed according to the manufacturer’s instructions using the SuperScript IV First-Strand Synthesis System (Invitrogen, Thermo Fisher Scientific). The subsequent multiplex PCRs (Even and Odd Mix) were performed using the V3 Primer Set in combination with the NEBNext Ultra II Q5 Master Mix (New England Biolabs, Frankfurt am Main, Germany). The only deviation from the original protocol was the reduction of the elongation time of the PCR to 45 seconds. The Even and Odd Mix PCR products were purified separately and their DNA concentration was determined using the Qubit™ dsDNA HS Assaykit (Thermo Fisher Scientific, Dreieich, Germany). The subsequent library preparation was executed with 25 ng of the Even and Odd PCR products. Finally, nanopore sequencing was performed using SQK-LSK109 chemistry on a 9.4.1 SpotON Flow Cell on the GridION system (Oxford Nanopore Technologies, Oxford, UK).

After passing sequence quality control, demultiplexing requiring barcodes at both ends of the reads was achieved using Porechop [15] and adapter trimming was achieved by the ARTIC pipeline [13] setting Wuhan-Hu-1 as reference strain for read mapping.

When the ARTIC pipeline failed or if the obtained consensus sequences were of too low quality or incomplete, the extracted RNA was again converted to cDNA using the SuperScript IV First-Strand Synthesis System (Invitrogen, Thermo Fisher Scientific). After performing the second strand synthesis (NEBNext Ultra II Non-Directional RNA Second Strand Synthesis Module, New England Biolabs), an Illumina library was generated using the NEBNext Ultra II FS DNA Library Prep Kit. Additionally, a target enrichment step was incorporated prior to sequencing on an Illumina MiSeq (Illumina Inc, Berlin, Germany). For this purpose, SARS-CoV-2 specific baits (myBaits Expert SARS-CoV-2, Arbor Sciences, BioCat, Heidelberg, Germany) were used according to the manufacturers’ instructions and captured libraries were sequenced using Sequencing V2 Reagent chemistry 300 cycles on a Micro Flow Cell (Illumina Inc, Berlin, Germany).

### Phylogenetic Analysis

For the enrichment pipeline, Illumina reads were mapped to the reference strain Wuhan/Hu-1/2019 (GenBank accession MN908947) using bwa v0.7.17 [16]. The obtained bam files were added as a separate read-group to the bam files generated by the ARTIC pipeline using SAMtools [17]. Follow-up steps of the ARTIC pipeline were performed manually using the combined bam files as input. Additionally, variants were called for the Illumina mapping using BCFtools v1.9-168 of the SAMtools package [17] and merged with the variant VCF files from the ARTIC pipeline, if necessary. All variations were screened against problematic_sites_sarsCov2.vcf for hompolasic sites or sequencing issues which have the potential to adversely affect phylogenetic and evolutionary inference [18]. Final genomes were submitted to GISAID to make them publicly available.

SNP and phylogenetic analyses were performed using a local installation of the nextstrain.org pipeline [5]. For this, strains listed in GISAID and belonging to the African subset (N=1203 as of June 22nd, 2020) were included in the initial analysis and further filtered to minimize redundancy and selected by relevance to possible travel and/or trade routes (N=73).

## Results

### Testing of Malian patient samples

Between the 3rd and 14th April 2020, 217 suspected cases (141 individuals living in Bamako and its surroundings and 76 passengers arriving from a return flight from Tunisia) were analyzed at the *Centre d’Infectiologie Charles Mérieux du Mali* for the presence of SARS-CoV-2 (Figure 1). A sample was only confirmed positive for a SARS-CoV-2 infection if the extracted RNA comprised both, the *E* gene and the *RdRp* gene. Via this approach, 38 patients (37/141 and 1/76) were diagnosed with COVID-19 representing 32,2% of all registered infections in Mali (N=118) during the given time course.

The age of the infected patients ranged from 2-82 years, while significantly more male (23/38) than female (12/38) individuals were tested positive for SARS-CoV-2.

**Figure 1.**
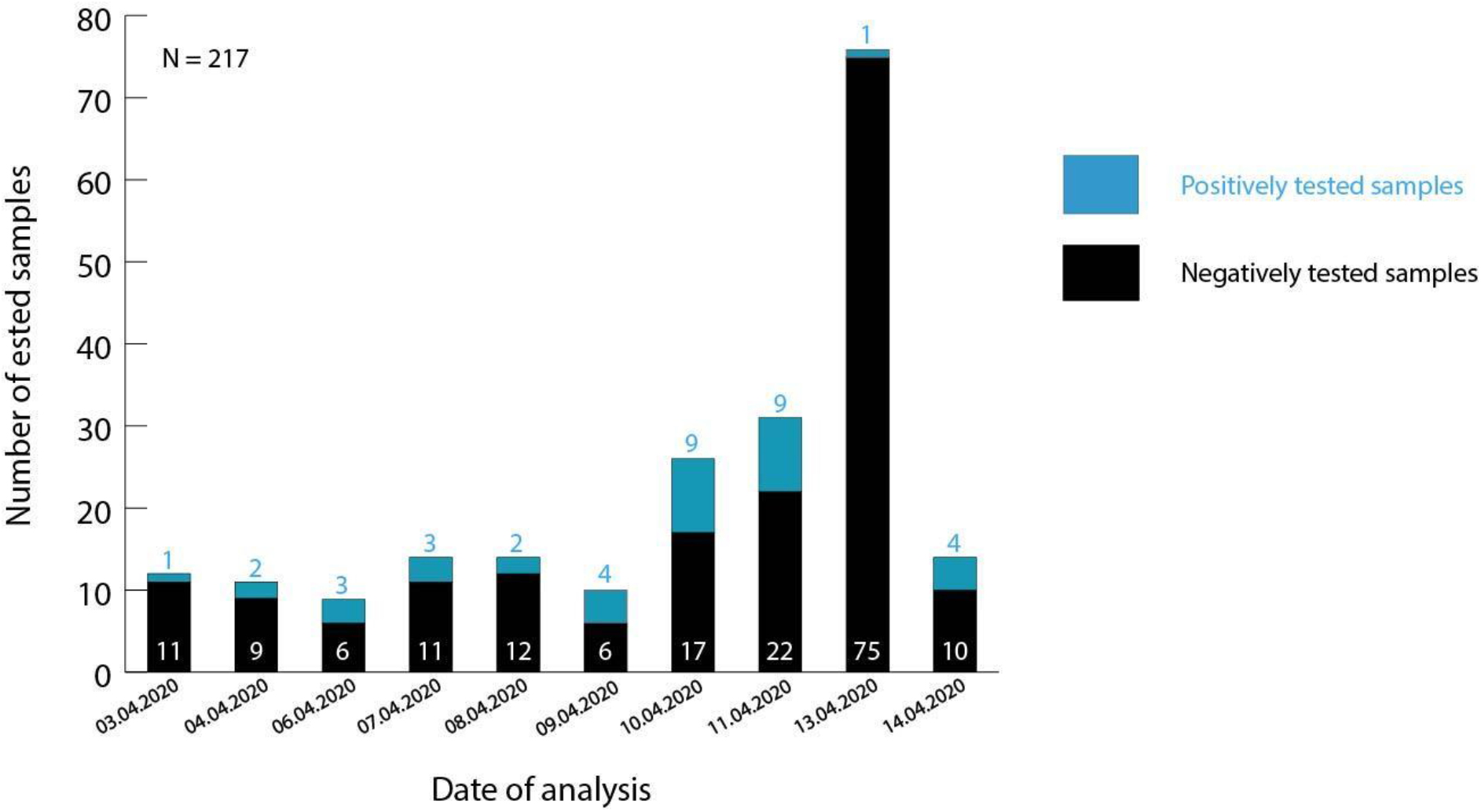
Number of positively confirmed samples analyzed at the *Centre d’Infectiologie Charles Mérieux du Mali* between 3rd and 14th April, 2020. Patient samples tested on the 13th of April represent screened passengers of a return flight from Tunisia. Residual samples reflect suspected cases from Bamako and its surrounding regions. Samples containing the *E* and *RdRp* gene were confirmed as positive.

Extracted RNA of positively tested patients was transferred to the Institute of Microbiology for whole genome sequencing analysis, as qualified sequencing capacities are locally not available and this method is used as an extension of the currently used diagnostic algorithms.

### Combination of short-read and long-read sequencing technologies resulted in the first 21 full length genomes of Mali

Ten full-length genome sequences could be successfully gained by using solely the ARTIC protocol and further 11 SARS-CoV-2 genomes could be obtained using an hybrid approach combining amplicons of the ARTIC protocol with the sequence reads from the enrichment workflow (see figure 2 and table 1). All genomes cover 29749 to 29858 nucleotides and reached a minimum sequencing depth of 40, thereby classifying themselves as high-quality genomes. Unfortunately, for 13 samples, even this advanced sequencing workflow resulted in only partially assembled genomes. Those isolates were neglected in our further analyses. It is worth noticing that a clear correlation between the Ct values of the *RdRp* gene and a successful sequencing was observed, showing that full-length genomes couldn’t be extracted from patient samples with Ct values higher than 32 (data not shown).

**Figure 2.**
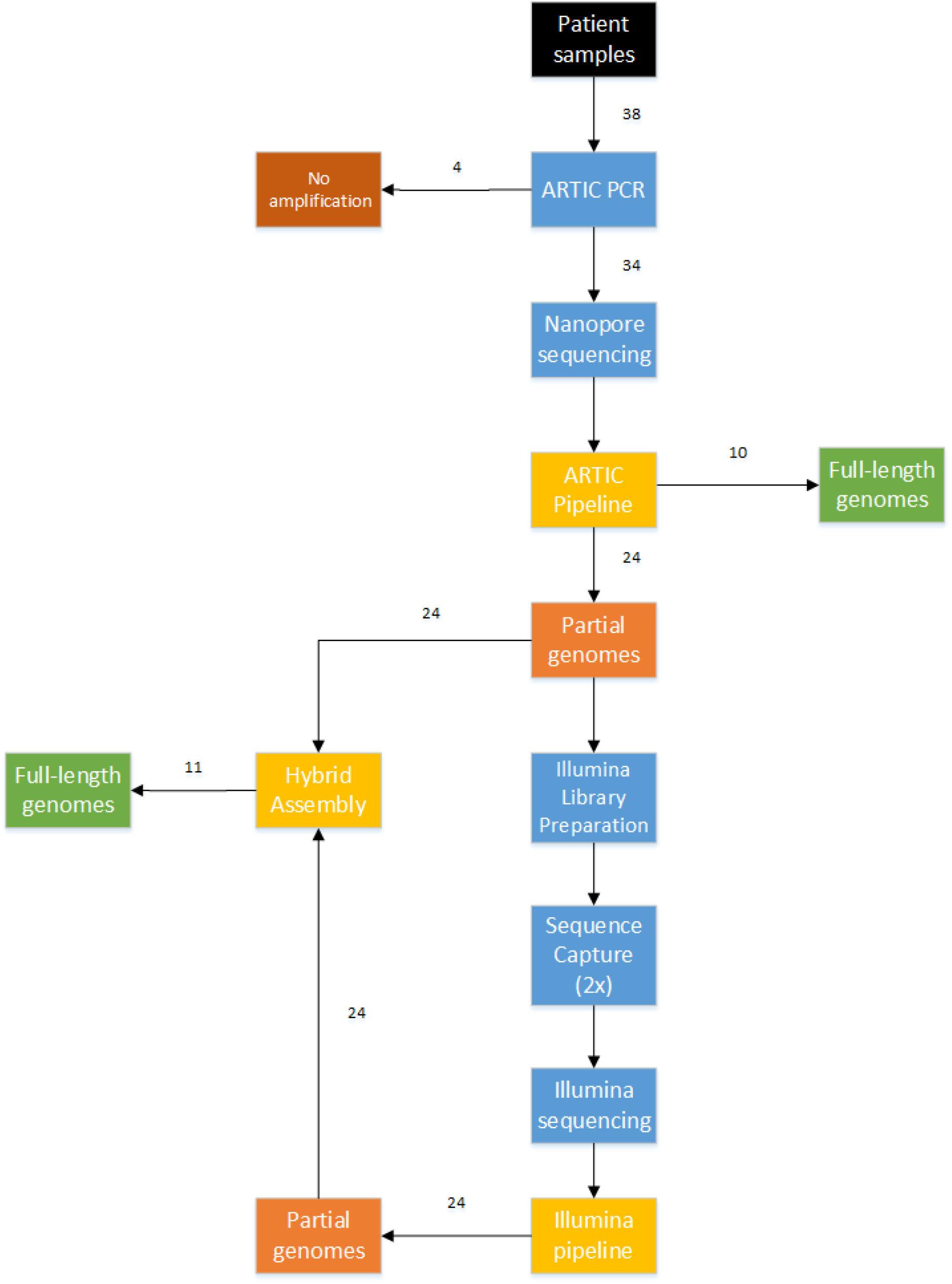
Workflow used to successfully sequence 21 out of 38 patient samples.

**Table 1.** Overview of patient information, sequencing results and used technology results of Malian samples (M = Male, F = Female, n.d. = not determined, A = ARTIC Protocol, H = Hybrid assembly of ARTIC protocol and bait-based enrichment using Illumina MiSeq, het = heterozygous, hp = highly homoplasic). Only non-synonymous mutations and deletions are listed.

| Sample ID | Patient<br>Sex,<br>Age | Location | AccNo<br>GISAID | Lineage<br>(Nextstrain<br>Clade) | Seq-Methods | Reference<br>covered<br>positions | SNPs (AA exchanges) |
| --- | --- | --- | --- | --- | --- | --- | --- |
| M002593 | M, 50-60 | Bamako | EPI_ISL487446 | A (19B) | A | 55 - 29835 | G11083T (ORF1a: L3606F, hp)<br>T28144C (ORF8:L84S)<br>G28878A (N:S202N)<br>C29585T (ORF10:P10S)<br>G29742A (3'UTR) |
| M002615 | M, 40-50 | Sévaré | EPI_ISL_4874 | A (19B) | H | 55 - 29863 | C25904T (ORF3a:S171L)<br>T28144C (ORF8:L84S)<br>G28878A (N:S202N)<br>G29742A (3'UTR) |
| M002616 | F, 40-50 | Bamako | EPI_ISL_487448 | A (19B) | H | 19 - 29776 | C14189T (ORF1b:A241V)<br>T28144C (ORF8:L84S)<br>G28878A (N:S202N)<br>G29742A (3'UTR) |
| M002618 | M, 60-70 | Bamako | EPI_ISL_487449 | A (19B) | H | 16 - 29871 | G581A (ORF1a:V106I)<br>A10323G(ORF1a:K3353R)<br>T28144C (ORF8:L84S)<br>G28878A (N:S202N)<br>G29742A (3'UTR) |
| M002644 | F, 50-60 | Bamako | EPI_ISL_487450 | A (19B) | A | 55 - 29835 | A14255G (ORF1b:K263R)<br>T28144C (ORF8:L84S)<br>G28878A (N:S202N)<br>G29742A (3'UTR) |
| M002659 | M, 20-30 | Bamako | EPI_ISL_487451 | A (19B) | H | 16 - 29872 | G11083T (ORF1a:L3606F, hp)<br>C26366T (E:A41V)<br>T28144C (ORF8:L84S)<br>G28878A (N:S202N)<br>G29742A (3'UTR) |
| M002663 | M, 20-30. | Bamako | EPI_ISL_487452 | A (19B) | H | 16 - 29860 | C10626T (ORF1a:A3454V)<br>C25904T (ORF3a:S171L)<br>C25936T (ORF3a:H182Y)<br>T28144C (ORF8:L84S)<br>G28878A (N:S202N)<br>G29742A (3'UTR) |
| M002667 | M, 20-30 | Bamako | EPI_ISL_487453 | B.1 (20C) | A | 55 - 29835 | C241T(5' UTR)<br>C1059T (ORF1a:T265I)<br>C14408T (ORF1b:P314L)<br>A23403G (S:D614G)<br>G25563T (ORF3a:Q57H) |
| M002672 | F, 50-60 | Bamako | EPI_ISL_487454 | A (19B) | A | 55 - 29826 | C241T(5' UTR)<br>685-693 gap (9 nt)<br>C1968T (ORF1a:T563I)<br>G28878A (N:S202N) |
| M002673 | F, 80-90 | Bamako | EPI_ISL_487455 | B.1 (20A) | A | 55 - 29835 | C14408T (ORF1b:P314L, het.) |
| M002675 | M, 60-70 | Bamako | EPI_ISL_487456 | A (19B) | H | 25 - 29859 | G11417T (ORF1a:V3718F)<br>C16658T (ORF1b:T1064I het.)<br>T28144C (ORF8:L84S)<br>G28878A (N:S202N)<br>G29742A (3'UTR) |
| M002698 | F, 60-70 | Bamako | EPI_ISL_487457 | A (19B) | H | 19 - 29800 | C25904T (ORF3a:S171L)<br>T28144C (ORF8:L84S)<br>G28878A (N:S202N) |
| M002700 | M, 60-70 | Bamako | EPI_ISL_487458 | B.1 (20A) | H | 126 - 29873 | G11128A (ORF1a:M3621I)<br>C14408T (ORF1b:P314L)<br>A23403G (S:D614G)<br>C25433T (ORF3a:T14I)<br>G25563T (ORF3a:Q57H)<br>T26038C (ORF3a:S216P)<br>29554-29602 gap (48 nt) |
| M002703 | F, 80-90. | Bamako | EPI_ISL_487459 | B.1 (20A) | H | 16 - 29839 | C241T(5' UTR)<br>C1968T (ORF1a:T568I)<br>C14408T (ORF1b:P314L)<br>G15743A (ORF1b:S759N)<br>C22290T (S:A243V)<br>A23403G (S:D614G) |
| M002704 | M, 50-60 | Bamako | EPI_ISL_487460 | A (19B) | H | 17 - 29870 | T28144C (ORF8:L84S)<br>G28878A (N:S202N)<br>G29742A (3'UTR) |
| M002707 | M, 60-70 | Bamako | EPI_ISL_487461 | A (19B) | A | 55 - 29835 | C16658T (ORF1b:T1064I)<br>G18973T (ORF1b:V1836F, het.)<br>T28144C (ORF8:L84S)<br>G28878A (N:S202N)<br>G29742A (3'UTR) |
| M002758 | M, 70-80 | Bamako | EPI_ISL_487462 | B.1 (20C) | A | 55 - 29835 | C241T(5' UTR)<br>C1059T (ORF1a:T265I)<br>G10974A (ORF1a S3570N)<br>G12292T (ORF1a M4009I)<br>C14408T (ORF1b:P314L)<br>A23403G (S:D614G)<br>G25563T (ORF3a:Q57H)<br>C26198T (ORF3a:T269M)<br>G29745A (3'UTR) |
| M002823 | M, 40-50 | Bamako | EPI_ISL_487463 | A (19B) | A | 55 - 29835 | C2189T (ORF1a:L642F)<br>G11417T (ORF1a:V3718F)<br>T28144C (ORF8:L84S)<br>G28878A (N:S202N) |
| M002824 | M, 0-10 | Bamako | EPI_ISL_487464 | B.1 (20C) | A | 55 - 29835 | C241T(5' UTR)<br>A963T (ORF1ab:E233V)<br>C1059T (ORF1a:T265I)<br>C11916T (ORF1a:S3884L)<br>C14408T (ORF1b:P314L)<br>A23403G (S:D614G)<br>G25563T (ORF3a:Q57H) |
| M002826 | F, 30-40 | Bamako | EPI_ISL_487465 | A (19B) | H | 19 - 29838 | G11417T (ORF1a:V3718F)<br>T28144C (ORF8:L84S)<br>G28878A (N:S202N)<br>G29742A (3'UTR) |
| M002830 | M, 80-90 | Bamako | EPI_ISL_487466 | B.1 (20A) | A | 55 - 29835 | C241T(5' UTR)<br>G8371T (ORF1a:Q2702H)<br>C12465T (ORF1a:A4067V)<br>C14408T (ORF1b:P314L)<br>G20102T (ORF1b:S2212I)<br>A23403G (S:D614G)<br>G25563T (ORF3a:Q57H) |

Within the 21 full-length genomes, we observed that depending on the sequencing approach, some nucleotides of the 5’UTR and 3’UTR regions were missing. This phenomenon was especially seen in sequences generated only by reads based on the ARTIC protocol. However, in genomes assembled from reads of the hybrid approach (combination of the ARTIC protocol with the Illumina sequencing), the 5’ UTR and 3’UTR were almost fully sequenced. Due to this fact, some indeed present SNPs in the 5’-UTR and 3’-UTR region might not have been called/observed, if only the ARTIC protocol was used to sequence the corresponding samples.

### Malian SARS-CoV-2 genomes can be assigned to two different lineages

Mapping of all Malian full-length genomes against the reference genome (Wuhan/Hu-1/2020) revealed SNPs in all sequences. Most of these are already known from other strains sequenced during this pandemic. Nineteen of the identified SNPs are synonymous, while 38 SNPs are non-synonymous and are hence listed in Table 1 with addition of those located in the UTR regions. According to the quantity and position of the SNPs, the Malian SARS-CoV-2 genomes were assigned to clades. At the moment, three diverging nomenclatures exist: the Pangolin Lineages based on the Pangolin (Phylogenetic Assignment of Named Global Outbreak LINeages) algorithm [19], the GISAID clades using the actual letters of the marker mutations [20] and the nextstrain clades which are defined by year and a letter code [21]. As the Pangolin Lineage and the Nextstrain Clade resulted in a fairly similar classification for the Malian SARS-CoV-2 genomes (see figure 3), and it is up to now unsure which nomenclature will endure, we will discuss our results for both systems.

**Figure 3:**
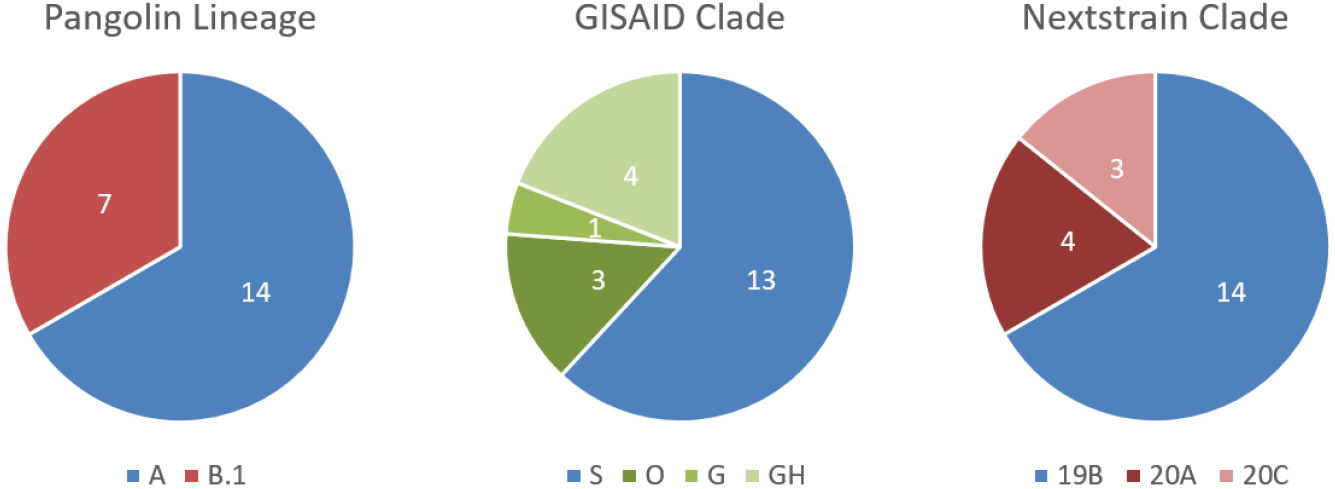
Sample assignment according to different nomenclatures used for SARS-CoV-2-classification.

Strains sequenced in this study can be assigned to the two major clades that occurred so far during this pandemic, namely A and B.1 according to the Pangolin lineage. While 14 genomes can be assigned to lineage A, which represents the lineage of the first observed strain of the pandemic in Wuhan (Wuhan/Hu-01/2020), 7 genomes belong to the currently worldwide fast spreading lineage B.1. According to the Nextstrain nomenclature, 14 strains belong to the 19B cluster, while the remaining 7 strains are split into the 20A (4) and 20C (3) clades. One of the main differences between the A and B.1 lineage and also the 19B and the 20* clades is the presence of the mutation D614G in the spike protein, which became dominant with the shift of the pandemic from China to Europe in February 2020. The further partition of the 20A cluster into the 20C clade is the gain of two further amino acid substitutions, namely T265I and Q57H (see also table 1).

Interestingly, all 14 genomes of lineage A (19B) cluster very closely to each other and are currently dominating this clade by number (Figure 4). Based on the reported sampling dates of the corresponding genome sequences, this clade was originally dominated by Asian samples (Country confidence Asia 81%, Europe 10%, Mali 4%, Oceania 2%), while recently, African and especially Malian genomes became more apparent (Country confidence Asia 58%, Europe 5%, Mali 36%, Oceania 0%).

**Figure 4:**
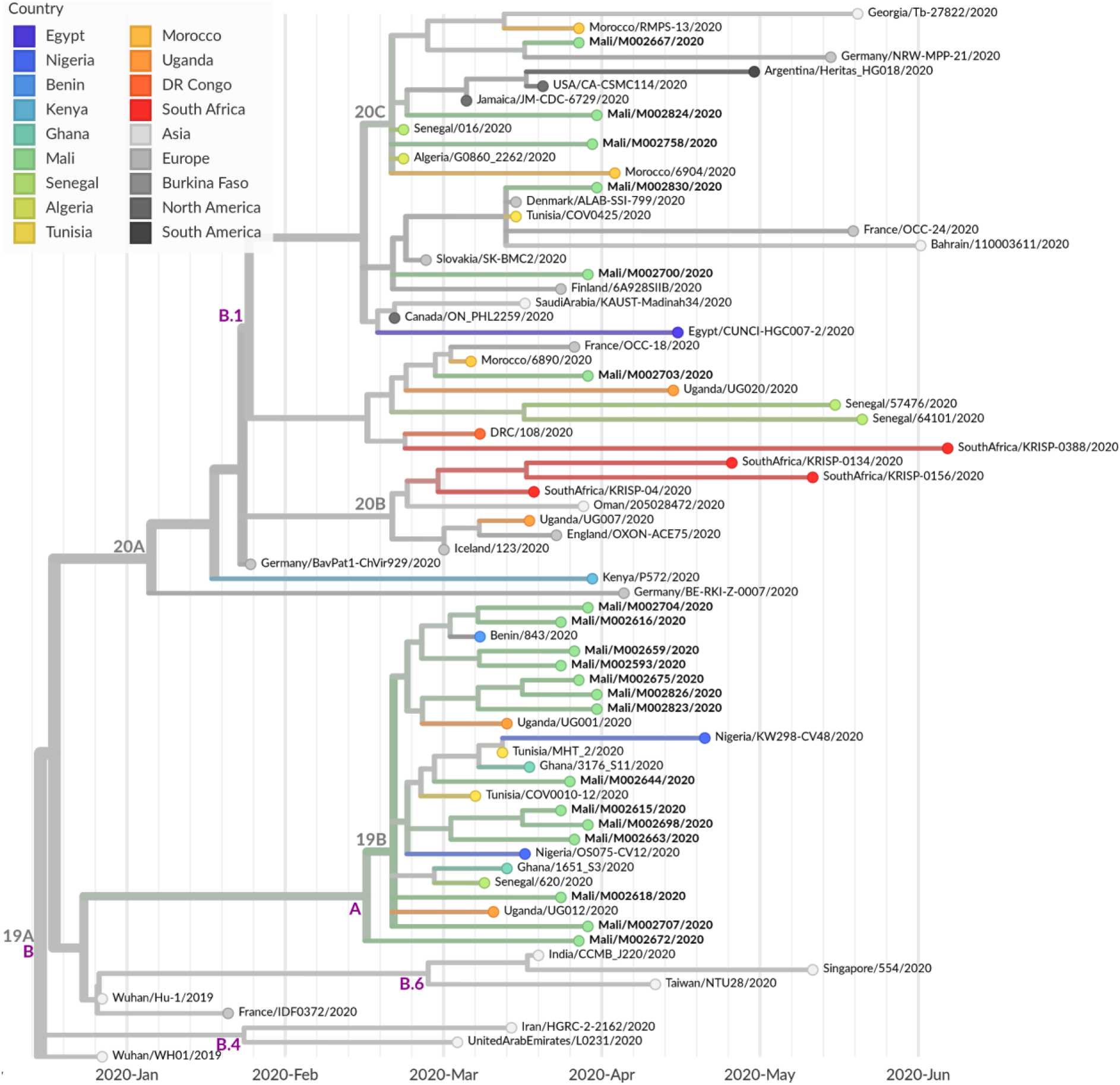
Maximum likelihood cladogram based on 47 representative genomes of African samples as well as 26 genetic closely related samples from Asia, Europe and North and South America. Nextstrain.org clade nomenclature is shown in gray and above branch nodes, Pangolin clade nomenclature is shown in violet and below branch nodes.

In contrast, the remaining 7 genomes are widely dispersed within the B.1 lineage (20A/C) (see figure 4), such as the genome sequence isolated from the positively tested Malian passenger (M00258), who returned from Tunisia. It also belongs to clade B.1, which might assume a possible infection outside of Mali. But as no information about symptome onset or Tunisia entry date has been available from this passenger, it cannot be excluded that he acquired the COVID-19 already in Mali prior his departure to Tunisia. Furthermore, nearest neighbour sequences to Malian genomes of this lineage are also locally widespread and originate from all over the world

### Special features of Malian genomes

Besides the division of the Malian genomes into the A (19B) and B (20A/C) clusters, two additional observations are worth mentioning.

The first one concerns sample M002672, belonging to the A (19B) lineage. Here, a 9 bp deletion at the nucleotide position 685, resulting in a 3 amino-acid deletion of the protein ORF1a, was detected. This microdeletion was already described in SARS-CoV-2 genomes derived from countries of the northern hemisphere like Iceland, Sweden, England, Wales, Canada and USA [22], but has, to our knowledge, not been detected in African samples so far. Unfortunately, a phenotypic function of this microdeletion remains to this day elusive.

The second observation was made in samples M002673 and M002707 at position 14408 and 18973, respectively. In both samples quasispecies, indicated by the simultaneous occurrence of at least two isolates, were detected at the described position. Table 1 lists only the dominant/more frequently occuring mutation. As the quasispecies were observed independent of the sequencing technology (Nanopore and Illumina), artificially introduced mutations via e.g. PCR can be excluded. The C14408T mutation, which leads to the P314L conversion in ORF1b is a known mutation and next to D614D another hallmark for the B (20*) cluster. This mutation seems to provoke a positive effect for the virus and can hence be regarded as directed selection. In contrast, the G18973T mutation, leading to the amino acid exchange V1836F, was only detected once before in a German cluster (example strain: Germany/NRW-MPP-24/2020), but did obviously not manifest. Nevertheless, we had the rare opportunity to witness the evolution of the virus within a human sample.

Regarding the variant screening we found only one variant in two strains, M002593 and M002659, which may confound evolutionary interpretations: G11083T. Position 11083 is a major homoplastic site in Orf1ab [23] which is geographically ubiquitous, and appears in strain M002659 both in the Illumina and MinION reads.

## Discussion

Up to now, only limited sequencing data of Northern Africa and the Sahel region is available compared to other regions of the world like America, Asia or Europe. Viral genomes spreading on the African continent are mainly accessible from states like South Africa [24], the Democratic Republic of Congo (DRC) and Kenya [25]. For the Sahel region, genomic information of SARS-CoV-2 is only provided by Nigeria [26] and Senegal [5,27], while data from countries like Mauritania, Niger, Burkina Faso, Chad, Eritrea and Sudan is still missing. With our full-length SARS-CoV-2 genomes originating from Mali, we want to contribute to the cohort of sequencing data from the Sahel region thereby enhancing the knowledge of the genomic diversity of COVID-19 infections in this country and helping the reconstruction of the geographic spread.

The Sahel region separates the northern African states (Morocco, Algeria, Tunisia, Libya, Egypt), which are historically and up to today linked to Europe, especially France, from the indigenous and distinct cultures from Central Africa. This unique geographical position makes the region exceptionally interesting for studying the origin of the COVID-19 pandemic. Our results show that two SARS-CoV-2 lineages, namely A and B.1 are circulating in Mali, with lineage A being causative for about ⅔ of the infections. Even though our findings might be biased due to the fact that mainly individuals from Bamako and its surroundings were sampled, the presence of the two lineages strongly suggests at least two different and independent introduction points of the SARS-CoV-2 infection in Mali.

Since the A lineage contains the initial emerging pandemic strain, as well as Asian genomes [19], we suggest a very early, but probably unrecognized onset of infections in Mali. Reasons that most likely encouraged the non-detection of the virus are on the one hand the fragile and overstretched health care system and on the other hand the transmission through patients with only mild symptoms being misdiagnosed or even not at all recognized. These listed reasons might also explain the relatively high mortality rate (5%) and the high estimated number of unreported cases in Mali.

In contrast, the B.1 lineage comprises sequences associated with the large Italian outbreak in February 2020 and represents to date (date 21.07.2020), together with its sublineage B.1.1, the worldwide prevalent virus clade [19]. The African continent, especially South Africa is dominated by lineage B (its origin still lying in Asia) and B.1 [24]. In Mali, only ⅓ of the patient samples were assigned to lineage B.1, thus giving rise to speculation that this viral lineage was imported at a later point in time. As the nearest neighbours of our Malian genomes belonging to the B.1 lineage and are geographically widely spread (Europe, USA, Canada, Northern Africa), we can exclude a single event as the origin of introduction of the B.1 lineage into Mali. More likely, multiple travel associated transfers from various countries have caused the spread of the B.1 lineage within Mali. Unfortunately, restrictive measures in order to contain the circulation within Mali are only possible to a limited extent since social distancing measures endanger vital income for survival and are hence not fully feasible.

To follow the three main rules of outbreak management, namely “test, trace and isolate”, the German Enhancement Initiative against Biological Threats in the G5 Sahel region have established the SARS-CoV-2 diagnostic at the *Centre d’Infectiologie Charles Mérieux du Mali* in Bamako, seeing the importance of strengthening and increasing testing capacities in Mali and Sahelian G5 partner countries. In doing so, we believe that a prompt identification of the source of infection will not only lead to a backtracking and isolation of individuals or groups but also to reasonable national measures to restrict or terminate the SARS-CoV-2 spread within the country and hence worldwide.

## Data Availability

All genomes are available at gisaid.org

## Acknowledgment

We gratefully acknowledge the Malian patients described in these case reports for giving written consents to the inclusion of material pertaining to themselves. We acknowledge the Institut National de la Santé Publique and the Direction Générale de la Santé for the sampling of the patients.

We acknowledge the authors from originating and submitting laboratories of sequence data provided by GISAID on which the analysis is based.

We gratefully thank Josua Zinner for excellent technical assistance during sequencing.

## Funding

This work was supported by the Enable and Enhance Initiative of the German Government ‘’Security cooperation on biological threats in Mali and G5-Sahel’’

(Ertüchtigungsinitiative der Deutschen Bundesregierung, OR12-370.43 ERT MLI/G5S) and Fondation Merieux, Lyon, France.

## References

1. Li, Q.; Guan, X.; Wu, P.; Wang, X.; Zhou, L.; Tong, Y.; Ren, R.; Leung, K.S.M.; Lau, E.H.Y.; Wong, J.Y.; et al. Early Transmission Dynamics in Wuhan, China, of Novel Coronavirus-Infected Pneumonia. N. Engl. J. Med. 2020, 382, 1199–1207, doi:10.1056/NEJMoa2001316.

2. Dong, E.; Du, H.; Gardner, L. An interactive web-based dashboard to track COVID-19 in real time. Lancet Infect. Dis. 2020, 20, 533–534, doi:10.1016/S1473-3099(20)30120-1.

3. World Health Organization Europe Coronavirus (COVID-19) | WHO | Regional Office for Europe Available online: https://www.euro.who.int/en/health-topics/health-emergencies/coronavirus-covid-19/novel-coronavirus-2019-ncov (accessed on Jul 10, 2020).

4. World Health Organization Africa Coronavirus (COVID-19) | WHO | Regional Office for Africa Available online: https://www.afro.who.int/health-topics/coronavirus-covid-19 (accessed on Jul 10, 2020).

5. Hadfield, J.; Megill, C.; Bell, S.M.; Huddleston, J.; Potter, B.; Callender, C.; Sagulenko, P.; Bedford, T.; Neher, R.A. Nextstrain: real-time tracking of pathogen evolution. Bioinformatics 2018, 34, 4121–4123, doi:10/gdkbqx.

6. Mali Government Sur les premieres cas de Coronavirus au Mali – Communique du gouvernement de la republique du Mali, 25. March 2020 2020.

7. The Food Crisis Prevention Network Food and nutrition crisis 2020 Available online: http://www.food-security.net/en/topic/food-and-nutrition-crisis-2020/ (accessed on Jul 10, 2020).

8. OECD coronavirus-west-africa - Sahel and West Africa Club Secretariat Available online: http://www.oecd.org/swac/coronavirus-west-africa/ (accessed on Jul 10, 2020).

9. Africa CDC - COVID-19 Daily Updates. Afr. CDC.

10. Corman, V.M.; Landt, O.; Kaiser, M.; Molenkamp, R.; Meijer, A.; Chu, D.K.; Bleicker, T.; Brünink, S.; Schneider, J.; Schmidt, M.L.; et al. Detection of 2019 novel coronavirus (2019-nCoV) by real-time RT-PCR. Euro Surveill. Bull. Eur. Sur Mal. Transm. Eur. Commun. Dis. Bull. 2020, 25, doi:10/ggjs7g.

11. COMMUNIQUE N°146 DU MINISTERE DE LA SANTE ET DES AFFAIRES SOCIALES SUR LE SUIVI DES ACTIONS DE PREVENTION ET DE RIPOSTE FACE A LA MALADIE A CORONAVIRUS. Available online: http://www.sante.gov.ml/index.php/actualites/communiques/item/3625-communique-n-146-du-ministere-de-la-sante-et-des-affaires-sociales-sur-le-suivi-des-actions-de-prevention-et-de-riposte-face-a-la-maladie-a-coronavirus (accessed on Jul 27, 2020).

12. Dreier, J.; Störmer, M.; Kleesiek, K. Use of Bacteriophage MS2 as an Internal Control in Viral Reverse Transcription-PCR Assays. J. Clin. Microbiol. 2005, 43, 4551–4557, doi:10.1128/JCM.43.9.4551-4557.2005.

13. Quick, J. nCoV-2019 sequencing protocol. 2020, doi:10/gg29d9.

14. Quick, J.; Lohman, N. hCoV-2019/nCoV-2019 Version 3 Amplicon Set Available online: https://artic.network/resources/ncov/ncov-amplicon-v3.pdf (accessed on Apr 28, 2020).

15. Wick, R.R. Porechop; 2017;

16. Li, H. Aligning sequence reads, clone sequences and assembly contigs with BWA-MEM. *ArXiv13033997 Q-Bio* 2013.

17. Li, H.; Handsaker, B.; Wysoker, A.; Fennell, T.; Ruan, J.; Homer, N.; Marth, G.; Abecasis, G.; Durbin, R. The Sequence Alignment/Map format and SAMtools. Bioinformatics 2009, 25, 2078–2079, doi:10.1093/bioinformatics/btp352.

18. De Maio, N.; Gozashti, L.; Turakhia, Y.; Walker, C.; Lanfear, R.; Corbett-Detig, R.; Goldman, N. Issues with SARS-CoV-2 sequencing data Available online: https://virological.org/t/issues-with-sars-cov-2-sequencing-data/473 (accessed on Jul 24, 2020).

19. Rambaut, A.; Holmes, E.C.; O’Toole, Ä.; Hill, V.; McCrone, J.T.; Ruis, C.; du Plessis, L.; Pybus, O.G. A dynamic nomenclature proposal for SARS-CoV-2 lineages to assist genomic epidemiology. Nat. Microbiol. 2020, 1-5, doi:10/gg47xd.

20. Freunde von GISAID e.V. Clade and lineage nomenclature aids in genomic epidemiology studies of active hCoV-19 viruses Available online: https://www.gisaid.org/references/statements-clarifications/clade-and-lineage-nomenclature-aids-in-genomic-epidemiology-of-active-hcov-19-viruses/ (accessed on Jul 22, 2020).

21. Year-letter Genetic Clade Naming for SARS-CoV-2 on Nextstain.org Available online: https://nextstrain.org//blog/2020-06-02-SARSCoV2-clade-naming (accessed on Jun 25, 2020).

22. Chrisman, B.; Paskov, K.; Stockham, N.; Jung, J.-Y.; Varma, M.; Washington, P.; Wall, D.P. Common Microdeletions in SARS-CoV-2 Sequences Available online: https://virological.org/t/common-microdeletions-in-sars-cov-2-sequences/485 (accessed on Jul 13, 2020).

23. van Dorp, L.; Acman, M.; Richard, D.; Shaw, L.P.; Ford, C.E.; Ormond, L.; Owen, C.J.; Pang, J.; Tan, C.C.S.; Boshier, F.A.T.; et al. Emergence of genomic diversity and recurrent mutations in SARS-CoV-2. Infect. Genet. Evol. 2020, 83, 104351, doi:10/ggvz4h.

24. Giandhari, J.; Pillay, S.; Wilkinson, E.; Tegally, H.; Sinayskiy, I.; Schuld, M.; Lourenço, J.; Chimukangara, B.; Lessells, R.J.; Moosa, Y.; et al. Early transmission of SARS-CoV-2 in South Africa: An epidemiological and phylogenetic report. *medRxiv* 2020, 2020.05.29.20116376, doi:10/gg5h53.

25. Githinji, G. Introduction and local transmission of SARS-CoV-2 cases in Kenya Available online: https://virological.org/t/introduction-and-local-transmission-of-sars-cov-2-cases-in-kenya/497 (accessed on Jul 10, 2020).

26. Onuliyi, P. SARS-CoV-2 Genomes from Nigeria Reveal Community Transmission, Multiple Virus Lineages and Spike Protein Mutation Associated with Higher Transmission and Pathogenicity Available online: https://virological.org/t/sars-cov-2-genomes-from-nigeria-reveal-community-transmission-multiple-virus-lineages-and-spike-protein-mutation-associated-with-higher-transmission-and-pathogenicity/494 (accessed on Jul 10, 2020).

27. Shu, Y.; McCauley, J. GISAID: Global initiative on sharing all influenza data – from vision to reality. Eurosurveillance 2017, 22, 30494, doi:10/gfwfbd.

